# Trends in Healthcare Costs among People Living with HIV in Ontario, Canada, 2003-2018: Results from a Population-Based Retrospective Cohort Study

**DOI:** 10.64898/2026.02.18.26346556

**Authors:** Min Xi, Asnake Yohannes Dumicho, Darrell H. S. Tan, Lisa Masucci, Ann N. Burchell, Alice Zwerling, Huiting Ma, Wei Zhang, OHTN Cohort Study Team, Sharmistha Mishra, Kednapa Thavorn

## Abstract

**Objective:** To quantify trends in annual mean healthcare costs per person living with HIV from 2003 to 2018 from a publicly funded healthcare system perspective.

**Design:** We conducted a retrospective population-based study using administrative health data in Ontario, Canada, including 25,842 people living with HIV diagnosed and entering care between 1992 and 2018. A nested cohort from the Ontario HIV Treatment Network Cohort Study (n=3,516) provided additional HIV-related characteristics.

**Methods:** Annual mean healthcare costs per person were estimated using a validated costing algorithm and inflated to 2025 Canadian dollars. Trends were examined overall and stratified by sociodemographic factors (age, sex, rurality, neighbourhood income, immigration status) and year of entry into HIV care. Within the nested cohort, trends were stratified by nadir CD4 count and any antiretroviral therapy use since diagnosis.

**Results:** Annual mean cost per person increased from $11,963 in 2003 to $16,721 in 2018. Medication costs remained the largest cost component throughout (47.4-61.7%) and closely mirrored overall trends. Higher annual mean costs were consistently observed among individuals diagnosed at older ages, lower-income neighbourhood residents, long-term Ontario residents (Canadian-born or immigrated before 1985), and individuals with nadir CD4<200cells/µL.

**Conclusion:** Medication expenditures continue to drive healthcare costs for people living with HIV. Cost containing strategies, including expanded generic substitution and strengthened price negotiation, may reduce costs without compromising outcomes. Persistent cost disparities highlight the need to address delayed treatment initiation and broader social determinants shaping HIV treatment access and sustained engagement in care.

## Introduction

Over the past three decades, more effective antiretroviral therapy (ART) regimens, earlier ART initiation, and increased availability of generics (i.e., less costly versions of brand name products) [1–6], supported by differentiated service delivery models designed to reduce structural barriers [7], have improved survival and quality of life for people living with HIV [8–10]. Within Canada’s publicly funded healthcare system, these improvements have reshaped HIV care and associated healthcare utilization. As HIV care evolves, understanding how healthcare costs have changed over time is essential for supporting sustainable and equitable care delivery.

Reliable, up-to-date estimates of healthcare costs for people living with HIV are necessary for health system planning, resource allocation, and economic evaluations of prevention and treatment strategies. However, such evidence remains limited, particularly in Canada. Existing studies often relied on self-reported data prone to recall bias or originated from non-Canadian jurisdictions with different healthcare systems [11–25]. Nine Canadian studies have reported costs following HIV diagnosis [26–34], six of which predate 2014 [27–29,31,33,34]. The only Ontario-based study used HIV incidence data from 1978 to 1996 and estimated the annual cost of living with HIV to be $788 million between 1997 and 2001 [27]. Recent work estimated a national cost burden of $2.1 billion in 2021 [30]. Analyses from Alberta showed declining per-person costs from 2014 to 2017 [26]. In Canada, higher costs have been consistently observed among people living with HIV with advanced disease at diagnosis (i.e., lower CD4 counts) [26,30] or with comorbidities [32]. However, many of these provincial and national estimates relied on or were extrapolated from single-centre data from Southern Alberta, leaving gaps in population-level evidence, especially cost disparities by sociodemographic or clinical characteristics [35–37].

To address this gap, we leveraged linked administrative and clinical datasets to estimate annual healthcare costs per person living with HIV in Ontario, Canada, between 2003 and 2018. We characterized overall cost trends and examined variation by sociodemographic and clinical characteristics to better understand how costs have evolved within a universal, publicly funded healthcare system.

## Methods

### Study design and setting

We conducted a population-based, retrospective, cohort study of people living with HIV in Ontario, Canada’s most populous province (∼14.7 million residents) [38]. The study is reported in accordance to the Strengthening the Reporting of Observational Studies in Epidemiology guidelines (Appendix A) [39]. Ontario’s publicly funded healthcare system covers medically necessary services, including physician visits, hospitalization, ambulatory care, emergency department visits, and continuing care. However, unlike the remainder of HIV care, prescription drug coverage, including ART, is not universal. Medications may be accessed through the Ontario Drug Benefit program for eligible groups (e.g., individuals aged <25 or ≥65 years, social assistance recipients, individuals with high drug costs relative to income), and through other government-sponsored programs or private insurance [40]. In Canada, 14.4% of HIV care is paid out-of-pocket and 11.9% by private insurance [41]. In the Ontario HIV Treatment Network Cohort Study (OCS), 66.6% of people living with HIV were covered by government-sponsored programs, 20.3% by private insurance, and 4.8% paid for medications out-of-pocket [40].

### Data sources

We linked administrative health datasets housed at ICES (formerly the Institute for Clinical Evaluative Sciences) with OCS data, an open, multi-site cohort of people living with HIV with annual questionnaires since 2007 [42]. ICES is an independent, non-profit research institute in Ontario, Canada, whose legal status under Ontario’s health information privacy law allows it to collect, house, and analyze administrative health data for Ontario residents with a valid health card, representing >99% of all Ontario residents [43].

### Study population

People diagnosed with HIV and in care were identified using a validated case-finding algorithm applied to the ICES HIV and OCS databases [44]. The algorithm required ≥3 HIV-related physician billing claims within 3 years (International Classification of Diseases 9^th^ and 10^th^ edition codes 042-044 and B20-B24); this algorithm demonstrated high validity (sensitivity: 96.2%, specificity: 99.6%) [44]. The diagnosis date was the earlier of the first HIV-related billing claim or the OCS-recorded diagnosis date.

We included individuals of all ages who were diagnosed with HIV and accessed care between April 1, 1992, and March 31, 2018, and had a valid Ontario health card. For OCS individuals, inclusion required written consent for ICES linkage and completion of at least one baseline questionnaire or interview between April 1, 1995 and March 31, 2018. Exclusion criteria included: invalid Ontario health card, age>105 years, missing value for sex, invalid or implausible birth date, or invalid death date.

We followed individuals until death (as recorded in the Registered Persons Database), loss to follow-up, or the end of the study (March 31, 2019), whichever occurred first. The accrual period ended on March 31, 2018, corresponding to the latest linkage between OCS and ICES data and to allow for at least one full year of follow-up for cost estimation.

### Covariates

Sociodemographic variables were measured at cohort entry and included: age, sex (Registered Persons Database), rurality and neighbourhood-level income quintile. Rural residence was defined as residing outside the commuting zone of a city with a population >10000 [45]. Income quintile was determined based on median household income relative to each individual’s neighbourhood in the Statistics Canada census year closest the individual’s year of cohort entry. Immigration status (Canadian-born or immigrated before 1985, longer term immigrant [greater than five years and immigrated after 1985], newcomer [within five years prior to cohort entry]) was derived from the Immigration, Refugee and Citizenship Canada Permanent Resident Database.

Clinical variables were limited to the nested OCS cohort and included ART use (at diagnosis or ever) and nadir CD4 count (lowest CD4 count anytime at or after cohort entry). We obtained healthcare use data from the Ontario Health Insurance Plan database, the Ontario Drug Benefit database, the National Ambulatory Care Reporting System, the Canadian Institute for Health Information Discharge Abstract Database, the Same Day Surgery database, and the Ontario Mental Health Reporting System. All datasets were linked using unique encoded identifiers and analyzed at ICES.

### Mean annual healthcare costs per person living with HIV

Our primary outcome was the mean annual direct healthcare costs per person, estimated from a publicly funded healthcare payer perspective (Ontario Ministry of Health). Costs were derived using a validated ICES costing algorithm, widely applied in previous studies [46–49]. The algorithm assigned costs to health encounters in the following categories: hospitalizations, emergency department visits, physician services, ambulatory care, continuing care (e.g., long-term care, rehabilitation, home care), and publicly funded medications. For hospital-based services, costs were estimated by multiplying the resource intensity weight by the cost per weighted case (or per weighted day, where applicable) [48]. Costs for services billed to the Ontario Health Insurance Plan, Ontario Drug Benefit, and home care were estimated by multiplying the number of services used by the unit cost of each service [48]. Medication costs were restricted to the costs of medications dispensed to individuals covered by the Ontario Drug Benefit program. All costs were reported as mean annual healthcare costs per person, stratified by category as recommended by Canada’s Drug Agency [50]. Costs were inflated to 2025 Canadian Dollars (CAD) [51].

### Statistical analysis

We described sociodemographic and HIV-related clinical characteristics at cohort entry using median and interquartile range (IQR) for continuous variables and counts and proportions for categorical variables. We used the analysis of variance test to compare mean age at diagnosis across HIV diagnosis periods. Annual mean costs were reported from 2003 to 2018, stratified by sex and costing category. We estimated the percentage change in mean costs overall and within pre-defined time periods (2003-2009, 2010-2014, and 2015-2018), reflecting changes in HIV care. Subgroup analyses were conducted to obtain sex-adjusted mean annual costs by sociodemographic and HIV-related clinical characteristics.

### Ethics

Section 45 of Ontario’s Personal Health Information Protection Act authorizes ICES to collect personal health information, without consent, for the purpose of analysis or compiling statistical information with respect to the management of, evaluation or monitoring of, the allocation of resources to or planning for all or part of the health system. Projects conducted under section 45 do not require review by a Research Ethics Board. Our project was conducted under section 45 and approved by ICES’ Privacy and Legal Office. Ethics approval for the OCS was obtained from each study site. Participants signed informed consent at the time of OCS enrolment.

## Results

### Study population

We identified 25,842 unique people living with HIV in Ontario between 2003 and 2018, including 25,808 from the ICES-HIV database and 3,516 from the OCS database (Fig. 1). Throughout the study period, 4,700 individuals (18.2%) died. The median age at cohort entry was 39.0 years (IQR: 31.0-46.0; Table 1). Individuals diagnosed before 2000 were older at cohort entry compared to those diagnosed in later years (p<0.001). At cohort entry, most were male (n=20,079, 77.7%), Canadian-born or long-term immigrants (n=20,275, 78.5%), urban residents (n=24,793, 95.9%) and over one-third (n=9,094, 35.2%) lived in the lowest income quintile neighbourhoods. Among OCS participants (n=3,516), 80.6% (n=2,835) had ever initiated ART following diagnosis. The median nadir CD4 count was 211 cells/µL (IQR: 90-330); nearly half (n=1,589, 45.2%) had nadir CD4<200 cells/µL.

**Fig. 1:**
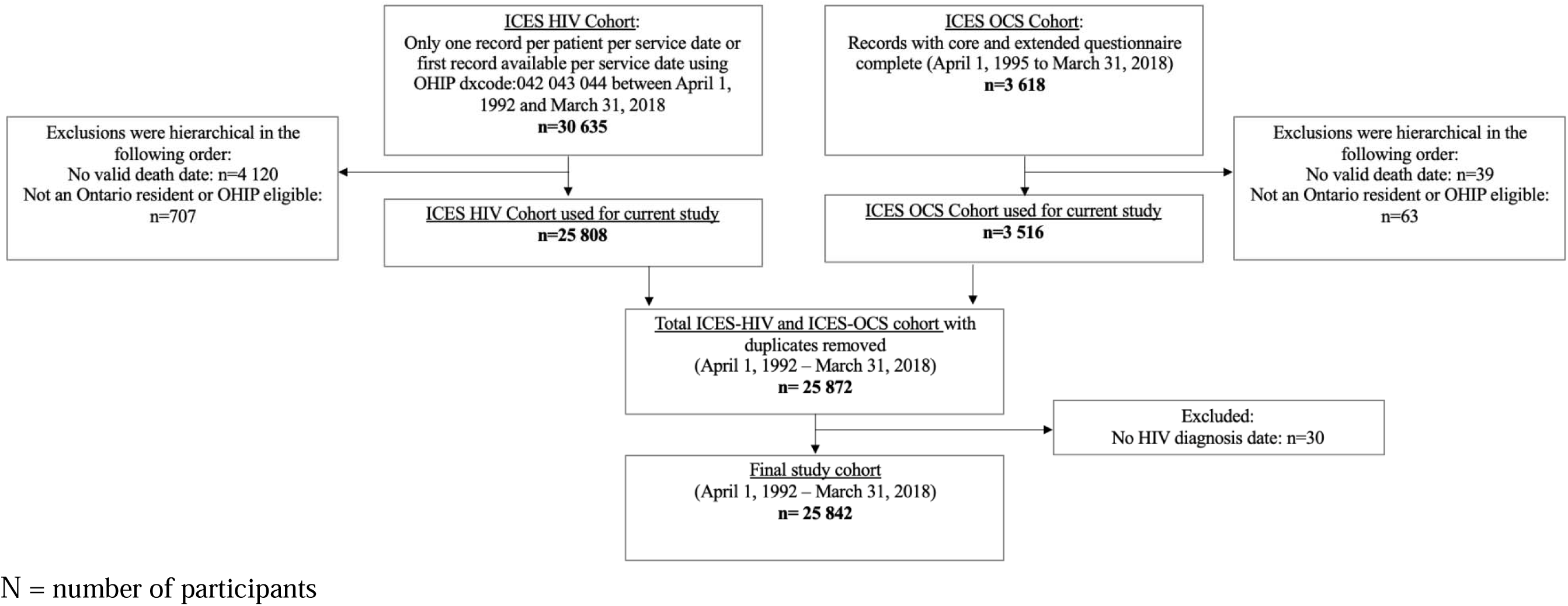
Participant flow diagram.

**Table 1.**
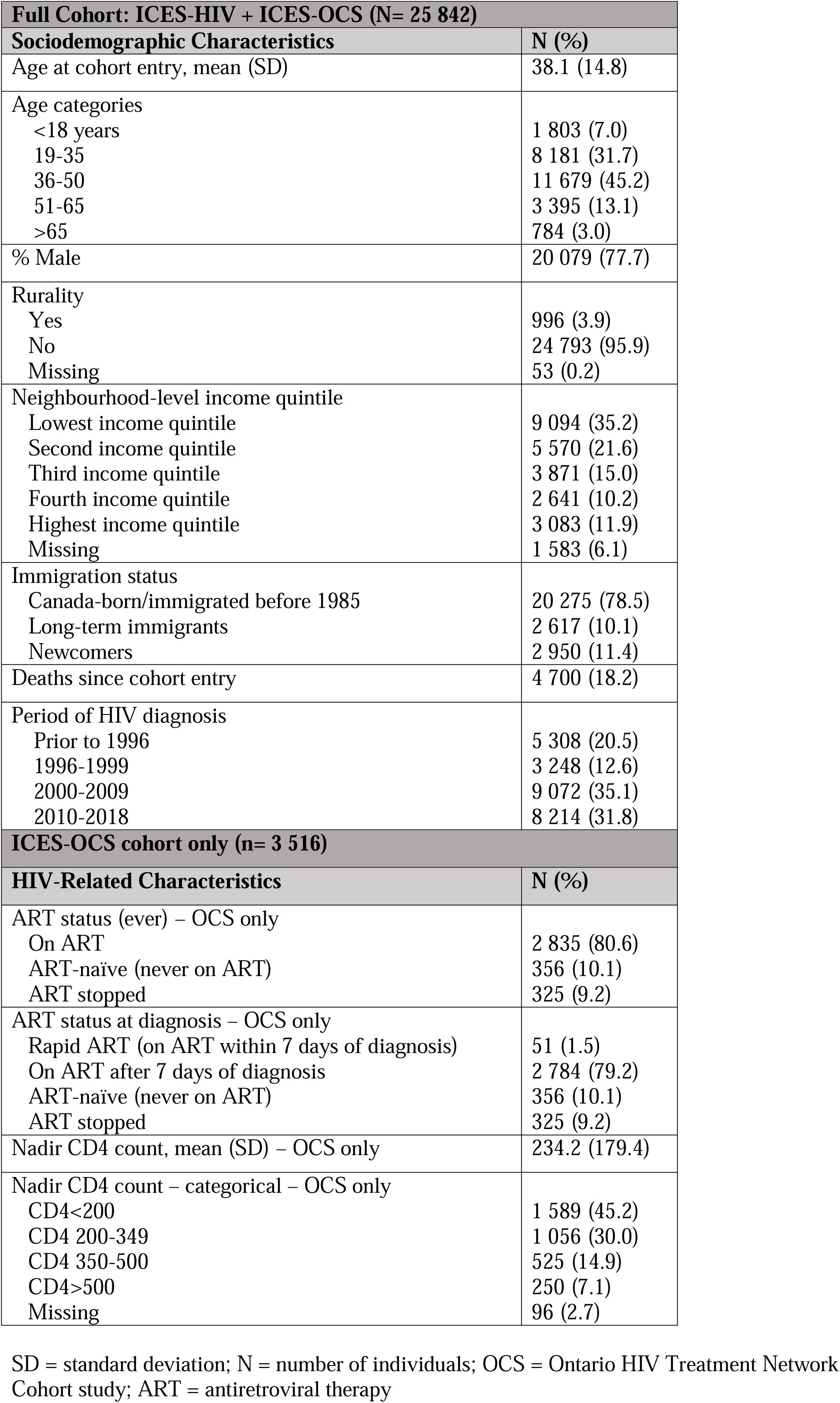
Participant characteristics at time of cohort entry.

| Full Cohort: ICES-HIV + ICES-OCS (N= 25 842) |  |
| --- | --- |
| Sociodemographic Characteristics | N (%) |
| Age at cohort entry, mean (SD) | 38.1 (14.8) |
| Age categories |  |
| <18 years | 1 803 (7.0) |
| 19-35 | 8 181 (31.7) |
| 36-50 | 11 679 (45.2) |
| 51-65 | 3 395 (13.1) |
| >65 | 784 (3.0) |
| % Male | 20 079 (77.7) |
| Rurality |  |
| Yes | 996 (3.9) |
| No | 24 793 (95.9) |
| Missing | 53 (0.2) |
| Neighbourhood-level income quintile |  |
| Lowest income quintile | 9 094 (35.2) |
| Second income quintile | 5 570 (21.6) |
| Third income quintile | 3 871 (15.0) |
| Fourth income quintile | 2 641 (10.2) |
| Highest income quintile | 3 083 (11.9) |
| Missing | 1 583 (6.1) |
| Immigration status |  |
| Canada-born/immigrated before 1985 | 20 275 (78.5) |
| Long-term immigrants | 2 617 (10.1) |
| Newcomers | 2 950 (11.4) |
| Deaths since cohort entry | 4 700 (18.2) |
| Period of HIV diagnosis |  |
| Prior to 1996 | 5 308 (20.5) |
| 1996-1999 | 3 248 (12.6) |
| 2000-2009 | 9 072 (35.1) |
| 2010-2018 | 8 214 (31.8) |
| ICES-OCS cohort only (n= 3 516) |  |
| HIV-Related Characteristics | N (%) |
| ART status (ever) – OCS only |  |
| On ART | 2 835 (80.6) |
| ART-naïve (never on ART) | 356 (10.1) |
| ART stopped | 325 (9.2) |
| ART status at diagnosis – OCS only |  |
| Rapid ART (on ART within 7 days of diagnosis) | 51 (1.5) |
| On ART after 7 days of diagnosis | 2 784 (79.2) |
| ART-naïve (never on ART) | 356 (10.1) |
| ART stopped | 325 (9.2) |
| Nadir CD4 count, mean (SD) – OCS only | 234.2 (179.4) |
| Nadir CD4 count – categorical – OCS only |  |
| CD4<200 | 1 589 (45.2) |
| CD4 200-349 | 1 056 (30.0) |
| CD4 350-500 | 525 (14.9) |
| CD4>500 | 250 (7.1) |
| Missing | 96 (2.7) |

SD = standard deviation; N = number of individuals; OCS = Ontario HIV Treatment Network Cohort study; ART = antiretroviral therapy

### Trends in mean annual healthcare costs

The mean annual healthcare cost increased by 39.8%, from $11,963 in 2003 to $16,721 in 2018. Costs and cost trends were similar for males and females (Fig. 2). Costs rose by 46.1% from $11,963 in 2003 to $17,473 in 2009. Costs remained relatively stable from 2010 to 2014, increasing modestly by 2.4% (2010: $17,164; 2014: $17,574), and declined by 6.6% from 2015 to 2018. Trends were similar for individuals diagnosed with HIV before 2015. However, individuals who were diagnosed with HIV between 2015 and 2018 experienced a 50.7% increase in costs between 2015 and 2017 (2015: $12,624; 2017: $19,027), followed by a 1.0% decline to $18,843 in 2018 (Fig. 3).

**Fig. 2:**
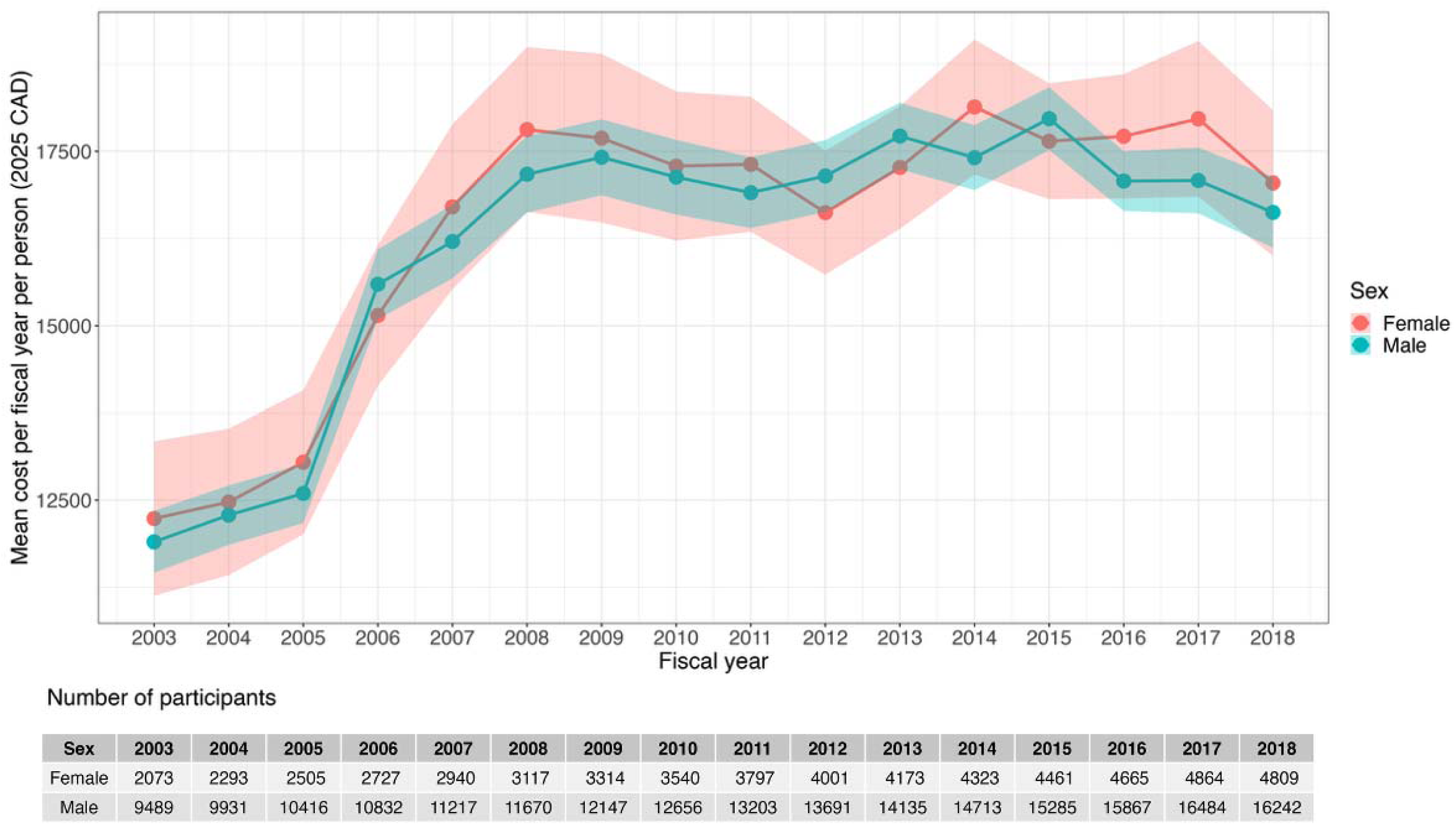
Mean annual direct healthcare costs per person by sex (2025 Canadian Dollars).

**Fig. 3:**
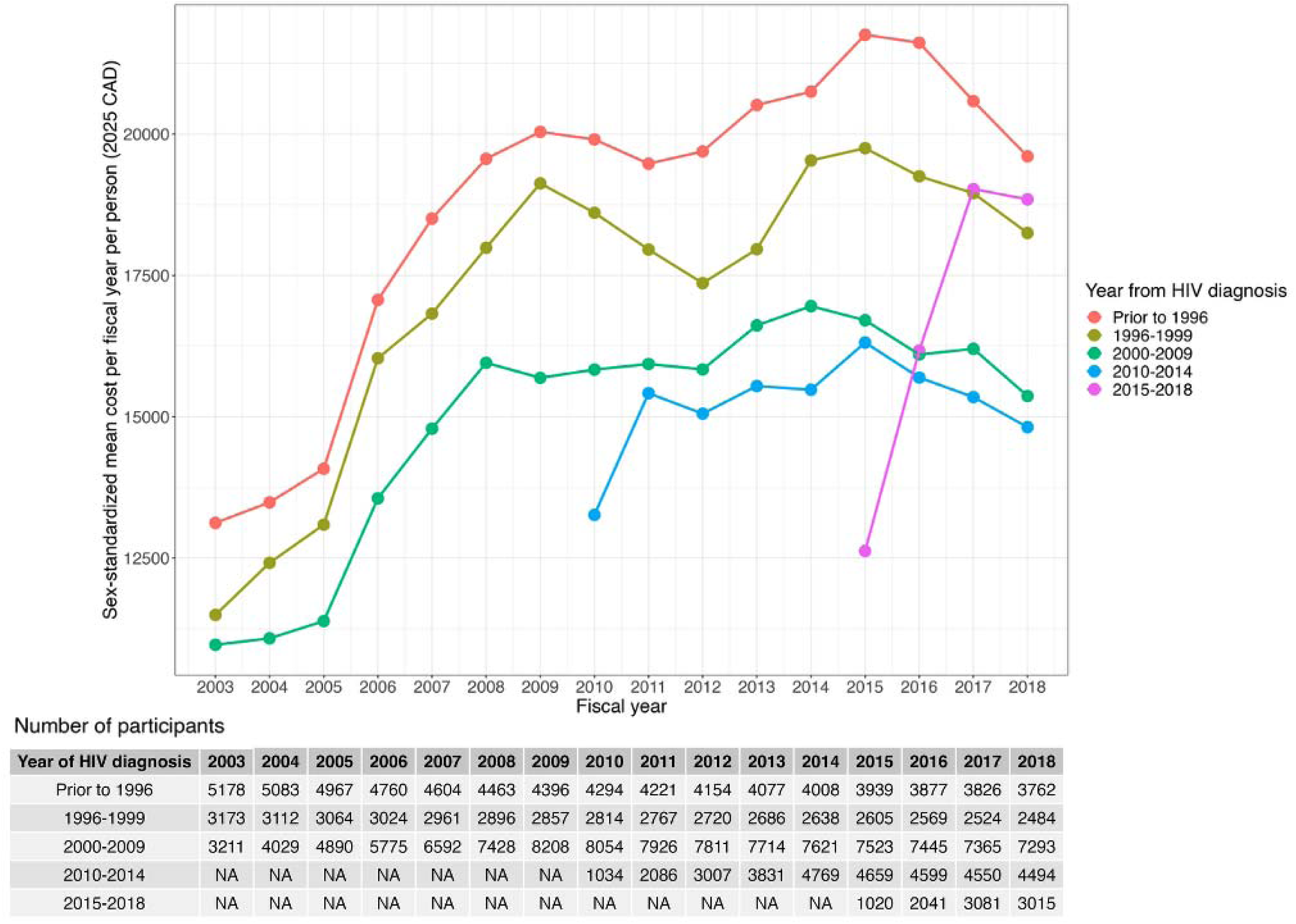
Mean annual direct healthcare costs per person by period of HIV diagnosis (2025 Canadian Dollars).

Medication costs consistently represented the largest share of total costs, ranging from 47.4% in 2006 to 61.7% in 2015 (Appendix B). Mean medication costs increased from $5,841 in 2003 to $11,038 in 2015, then declined to $9,677 in 2018. Hospitalization and physician services were the next highest contributors. Mean hospitalization costs rose from $2,410 in 2003 to $3,210 in 2009, then declined to $2,369 in 2015 before increasing to $2,826 in 2018. Mean physician costs declined from $1,898 in 2003 to $1,675 in 2018.

### Trends in mean annual healthcare costs by sociodemographic characteristics

Cost trajectories by sex, age (except for individuals aged ≤18 years and 19-35 years), rurality, immigration status, and neighbourhood-level income followed similar patterns, with a steep increase in mean annual healthcare costs from 2003 to 2009, followed by more stable or moderate increase in costs between 2010 and 2014, and a decline between 2015 and 2018 (Fig. 2; Appendices C-F). Notably, we observed a 17.6% decline in costs among newcomers from $12,235 in 2003 to $10,086 in 2005, before increasing to $14,728 in 2009. We found that older adults (>65 years), Canadian-born/long-term immigrants, and residents of the lowest-income neighborhoods consistently incurred higher mean annual costs compared to younger adults, newcomers, and those in higher-income neighborhoods.

### Trends in healthcare costs by clinical characteristics

Trends by nadir CD4 count mirrored overall patterns (Fig. 4). Costs across the nadir CD4 count categories increased by 47.0 to 82.8% from 2003 to 2009 and decreased by 4.2 to 22.6% between 2015 and 2018. Participants with low nadir CD4 count (<200 cells/µL) consistently experienced higher costs compared to participants in higher nadir CD4 categories. ART users consistently incurred higher costs compared to ART-naïve individuals except in 2008 (Appendix G). Among participants who were ART-naïve or discontinued ART, hospitalization costs accounted for a larger proportion of total costs compared to ART users (Appendix H). Conversely, ART users incurred higher medication costs (55.5%-65.9%) than ART-naïve participants (51.0%-53.4%; Appendix H).

**Fig. 4:**
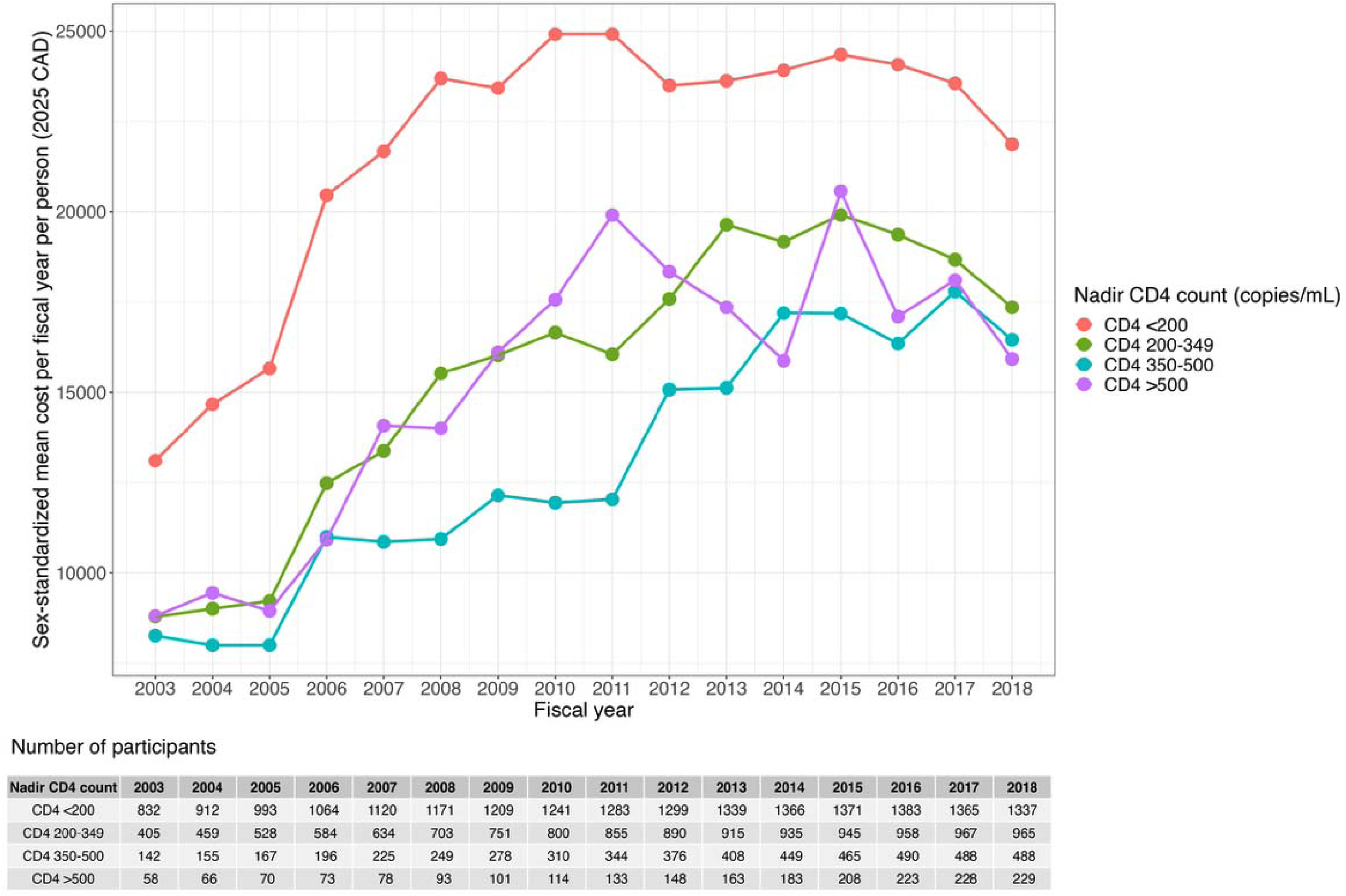
Mean annual direct healthcare costs per person by nadir CD4 count since HIV diagnosis (2025 Canadian Dollars).

## Discussion

In this large population-based cohort study, mean annual healthcare costs per person living with HIV in Ontario increased from 2003 until 2015 then declined through 2018. Medication costs consistently contributed to half or more of healthcare costs. Persistent disparities were evident, with older adults, individuals with low nadir CD4 counts, and people residing in socioeconomically disadvantaged neighbourhoods consistently incurring higher healthcare costs.

Our findings mirror previously reported Canadian and international HIV-related healthcare cost trends. In Alberta, Canada, mean annual healthcare costs rose 30% between 2006 and 2014, before declining 16% from 2014 to 2017 [26]. Globally, HIV spending across 188 countries increased from 2000 to 2013 before declining modestly from 2013 to 2015 [41]. These trends likely reflect shifts in HIV incidence and treatment practices over time. Given that costs for infectious diseases are typically highest immediately following diagnosis and near the end of life [52], rising number of new HIV diagnoses and linkage to care in Ontario from 2000 to 2015 likely contributed to higher expenditures in earlier years [53]. The rise in costs in the 2000s also reflect the uptake of more effective, newer, and more expensive branded ART regimens [54,55]. For example, Triumeq^TM^ (dolutegravir/abacavir/lamivudine) was introduced in Ontario in 2014 and was approximately 36-68% more expensive than existing regimens [56]. The subsequent decline in costs between 2015 and 2018 may be explained by the expanded availability and use of generic ART [26] and stabilization in linkage to care (87-88% from 2015 to 2018) and ART uptake (81-85% from 2015 to 2018) in Ontario [53].

Consistent with previous studies, ART and other medications were the main healthcare cost drivers in our study, accounting for 48 to 62% of mean annual costs. This range is slightly lower than estimates from other jurisdictions (60 to 79%) [15,26,57,58], potentially reflecting variation in ART pricing, public drug coverage across jurisdictions, and time periods [59]. In Ontario, approximately 25% of people living with HIV rely on private insurance or out-of-pocket payments for ART [40]. Because these costs are not captured in administrative data, our estimates represent a conservative assessment of medication-related costs.

Despite being the main cost driver, ART remains a high-value investment. Extensive evidence demonstrates that ART substantially improves health outcomes and quality of life [60,61] and prevents onward HIV transmission through sustained viral suppression [62–69]. Early ART initiation irrespective of CD4 count reduces serious AIDS- and non-AIDS-related events [70], which may lower future HIV care costs. Hence, ART spending should be interpreted not as inefficiency but as an essential driver of the population-level health gains observed over the past two decades. Optimizing ART expenditures remains critical for long-term system sustainability. Price negotiations, formulary management, and expanded use of generics offer opportunities for cost containment without compromising clinical outcomes. Modelling from the United Kingdom suggested that generic ART substitution, priced at 50 to 90% of branded ART, could reduce lifetime treatment costs by 42 to 69%, assuming generics of most current ART become available in 2027 and generics of newer ART become available in 2033 [71]. Similar generic substitution strategies in Canada [72], could yield significant savings that could be reinvested into priority areas such as increased HIV pre-exposure prophylaxis use, earlier diagnosis, improved linkage to care, expanded mental health and social supports, and addressing persistent inequities in HIV outcomes.

Our analyses emphasize persistent disparities in healthcare utilization and costs among people living with HIV across sociodemographic and clinical subgroups. Higher costs among older adults likely reflect the greater multimorbidity associated with ageing with HIV, resulting in longer hospital stays, specialist visits, and polypharmacy [32]. Age-related differences in drug coverage may further amplify this pattern. In Ontario, medication costs for individuals 65 years or older are publicly covered and fully captured in our estimates, whereas younger adults (25-64) often rely on a mix of public, private, and out-of-pocket coverage, leading to partial underestimation of their medication expenditures. Similar patterns have been reported in Alberta and Quebec, Canada, where costs among people living with HIV aged 50 years or older or with at least one comorbidity were 1.2 [34] and 2.7-fold [32] higher, respectively, compared to younger people living with HIV and matched controls.

Elevated costs observed among individuals diagnosed in earlier years and Canadian-born people living with HIV, or those who immigrated before 1985 may be partly attributable to the older age profile of these groups. Lower healthcare costs among newcomers and long-term immigrants may reflect the healthy immigrant effect (i.e., immigration screening process selects healthier individuals) [73–75]. Lower costs may also reflect the underuse of health services due to access barriers, under-detection of health conditions, limited system navigation support, or discrimination [76]. The socioeconomic gradient in costs aligns with established associations between structural disadvantage, higher HIV incidence, and greater comorbidity burden [77]. People living with HIV in lower-income Ontario neighbourhoods are more likely to qualify for public drug programs, meaning their ART costs are fully captured in administrative data. In contrast, medication costs for higher-income individuals are often covered by private insurance or paid out-of-pocket and therefore are not included in our estimates, resulting in an underestimation of their healthcare costs. These findings underscore the need for strategies that address the structural and access barriers driving cost disparities, including improving equitable access to screening, comorbidity management, and culturally safe care.

Similar to previous studies, we observed consistently higher costs among people living with HIV with a nadir CD4 count below 200 cells/µL compared to higher nadir CD4 count categories [26,33,78]. CD4 counts less than 200 cells/µL often mark serious health implications that require more intense monitoring, clinical management, and healthcare use, contributing to higher healthcare costs [79]. As CD4 counts are typically lower at diagnosis and rise following ART initiation [80], our findings emphasize the link between delayed diagnosis and ART initiation, advanced disease, and higher expenditures. Further research is needed to understand costs attributable to delayed versus early diagnoses and linkage to care.

Our study had several strengths. Using provincial administrative health data, we assembled a large, population-based cohort of people living with HIV in Ontario and estimated their longitudinal healthcare costs over 16 years. This is the first Ontario-specific cost study since 1998, addressing a major evidence gap and providing detailed cost estimates following HIV diagnosis within a universal healthcare system.

Nevertheless, several limitations should be considered. First, costs were estimated from the Ontario public payer perspective. Populations not fully covered under Ontario’s universal healthcare plan and/or individuals who face barriers to accessing care were not included in our analyses [81]. Given the perspective of our analyses, out-of-pocket expenditures, private insurance payments, and broader societal costs (e.g., productivity losses), were not included. These exclusions led to an underestimation of the mean annual healthcare cost per person living with HIV, particularly medication costs. Future research assessing costs from a societal perspective would provide a more comprehensive understanding of the economic impact of HIV. Second, given the secondary analysis of administrative health data, some covariates of interest were unavailable or limited to OCS participants (e.g., gender, race, sexual orientation, CD4 counts, ART use), which may not be representative of people living with HIV in Ontario. For most OCS sites, CD4 counts and ART use were extracted from clinical charts every six months [42], introducing potential measurement error that may have minimized cost differences between nadir CD4 count and ART use groups. Third, due to data linkage restrictions, we were unable to capture changes in costs beyond March 2019. Finally, our analyses were descriptive and did not reflect HIV-attributable costs. Subgroup analyses were based on characteristics at cohort entry and did not account for the time-varying nature of these characteristics. Future work using longitudinal modelling and comparative designs would help clarify the causal pathways underlying cost trajectories and allow estimation of the net healthcare costs attributable to HIV.

Our findings provide new evidence to guide cost projections, budget allocation, and economic evaluations of HIV prevention interventions in publicly funded healthcare systems. These estimates can help policymakers prioritize strategies that promote equity and sustainability in publicly funded healthcare systems. Our findings suggest that healthcare costs following an HIV diagnosis closely track HIV incidence and ART availability, with ART remaining the main cost driver. As new HIV diagnoses and related costs rise in Canada and similar contexts, strengthening drug price negotiation and expanding the use of generics may help curb spending while preserving health outcomes. Savings generated through these strategies could be reinvested in programs that strengthen HIV care and reduce preventable health system utilization.

Given an ageing population and ongoing disproportionate rates of new HIV diagnoses among socioeconomically disadvantaged groups [82–84], equity-informed programs are needed to ensure treatment and care benefits reach all people living with HIV and reduce downstream costs. Potential strategies include improving healthcare access for newcomers, expanding integrated and multidisciplinary models of care that address multimorbidity, expanded geriatric 1HIV services, and implementing interventions that address social and structural barriers to HIV testing, treatment, and care [85–87].

## Conclusion

Trends in healthcare costs following an HIV diagnosis in our study offer globally relevant insights. Negotiating lower prices for newer ART drugs and expanding generic drug use can help sustain health benefits while reducing costs. Our findings also reinforce the importance of early diagnosis, prompt ART initiation, and the need for community-based, culturally tailored, integrated models of care.

## Acknowledgements

We gratefully acknowledge the support of Dr Abigail Kroch, the REACH Team, Anan Bader Eddeen, Meltem Tuna, our community advisory board (LMJ Proulx, P Cupido), and volunteers and support teams of the Ontario HIV Treatment Network (OHTN) Cohort Study (OCS). We gratefully acknowledge all of the people living with HIV who volunteer to participate in the OHTN Cohort Study. We also acknowledge the work and support of OCS Governance Committee (Aaron Bowerman, Adrian Betts, Barry Adam, Cornel Gray, Dane Record, Mary Ndung’u, Michael Wilson, Rodney Rousseau, Ruth Cameron, YY Chen) OCS Scientific Steering Committee (Anita Benoit, Ann Burchell, Barry Adam, Curtis Cooper, David Brennan, Kelly O’Brien, Lance Mcready, Lawrence Mbuagbaw, Mona Loutfy, Pierre Giguere, Sean Hillier, Sergio Rueda (Chair), and Trevor Hart) and Indigenous Data Governance Circle (Meghan Young, Randy Jackson, Trevor Stratton). The OHTN also acknowledges the work of past Governance Committee and Scientific Steering Committee members.

We thank all interviewers, data collectors, research associates, coordinators, nurses, and physicians who provide support for data collection. The authors wish to thank OCS staff for data management, IT support, and study coordination: Chigozie Ugwu, Lucia Light, Mustafa Karacam, Nahid Qureshi, Tsegaye Bekele. The OHTN Cohort Study is supported by the Ontario Ministry of Health.

This study was supported by ICES, which is funded by an annual grant from the Ontario Ministry of Health (MOH) and the Ministry of Long-Term Care (MLTC). This study also received funding from the Canadian Institutes of Health Research (CIHR; 416186). This document used data adapted from the Statistics Canada Postal Code^OM^ Conversion File, which is based on data licensed from Canada Post Corporation, and/or data adapted from the Ontario Ministry of Health Postal Code Conversion File, which contains data copied under license from Canada Post Corporation and Statistics Canada. Parts of this material are based on data and/or information compiled and provided by and/or adapted from: Ontario Ministry of Health, CIHI, Statistics Canada (Census; current to June 3, 2022), Ontario Health, Johns Hopkins (ACG® System Version 10), IQVIA Solutions Inc., Immigration, Refugees and Citizenship Canada (IRCC; current to June 3, 2022). We thank IQVIA Solutions Canada Inc. for use of their Drug Information File. Parts of this material are based on data and information compiled and provided by the Ontario Ministry of Health. We thank the Toronto Community Health Profiles Partnership for providing access to the Ontario Marginalization Index. The analyses, conclusions, opinions and statements expressed herein are solely those of the authors and do not reflect those of the funding or data sources; no endorsement is intended or should be inferred.

## Conflicts of Interest

### Data availability

The dataset from this study is held securely in coded form at ICES. While legal data sharing agreements between ICES and data providers (e.g., healthcare organizations and government) prohibit ICES from making the dataset publicly available, access may be granted to those who meet pre-specified criteria for confidential access, available at www.ices.on.ca/DAS. The full dataset creation plan and underlying analytic code are available from the authors upon request, understanding that the computer programs may rely upon coding templates or macros that are unique to ICES and are therefore either inaccessible or may require modification.

