## Appendix B for "Trends in Healthcare Costs among People Living with HIV in Ontario, Canada, 2003-2018: Results from a Population-Based Retrospective Cohort Study"

**Appendix B.1:** Proportion of mean cost per person per year by costing category.

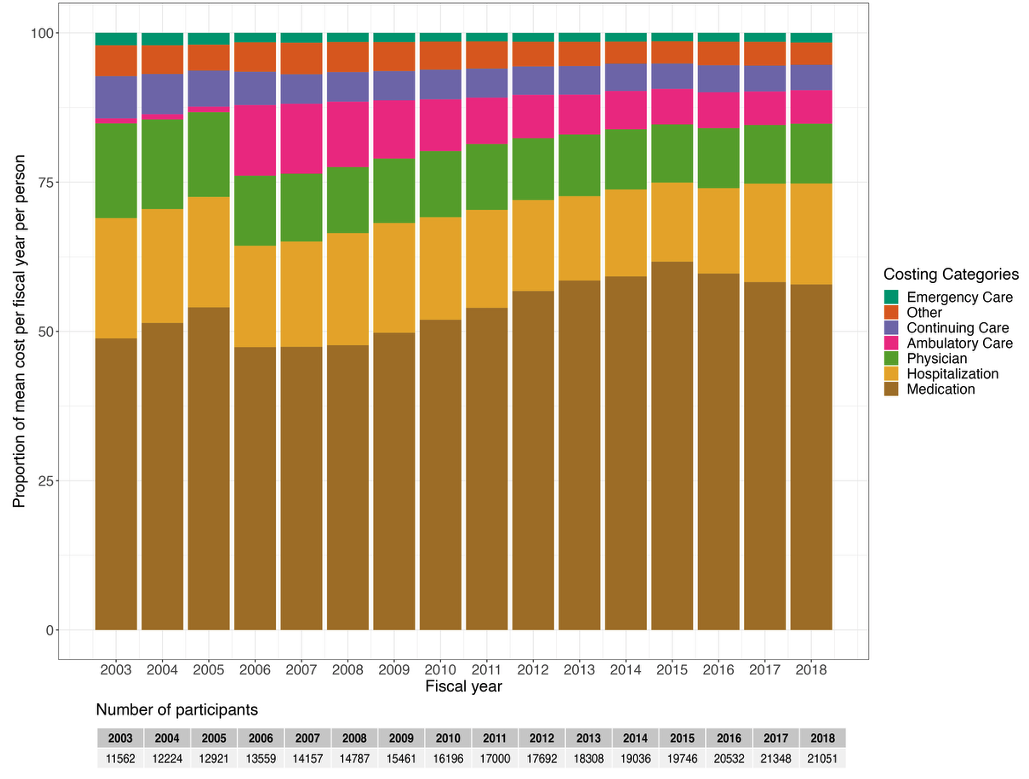

**Appendix B.2:** Mean annual healthcare cost per person per year by costing category (2025 Canadian dollars).

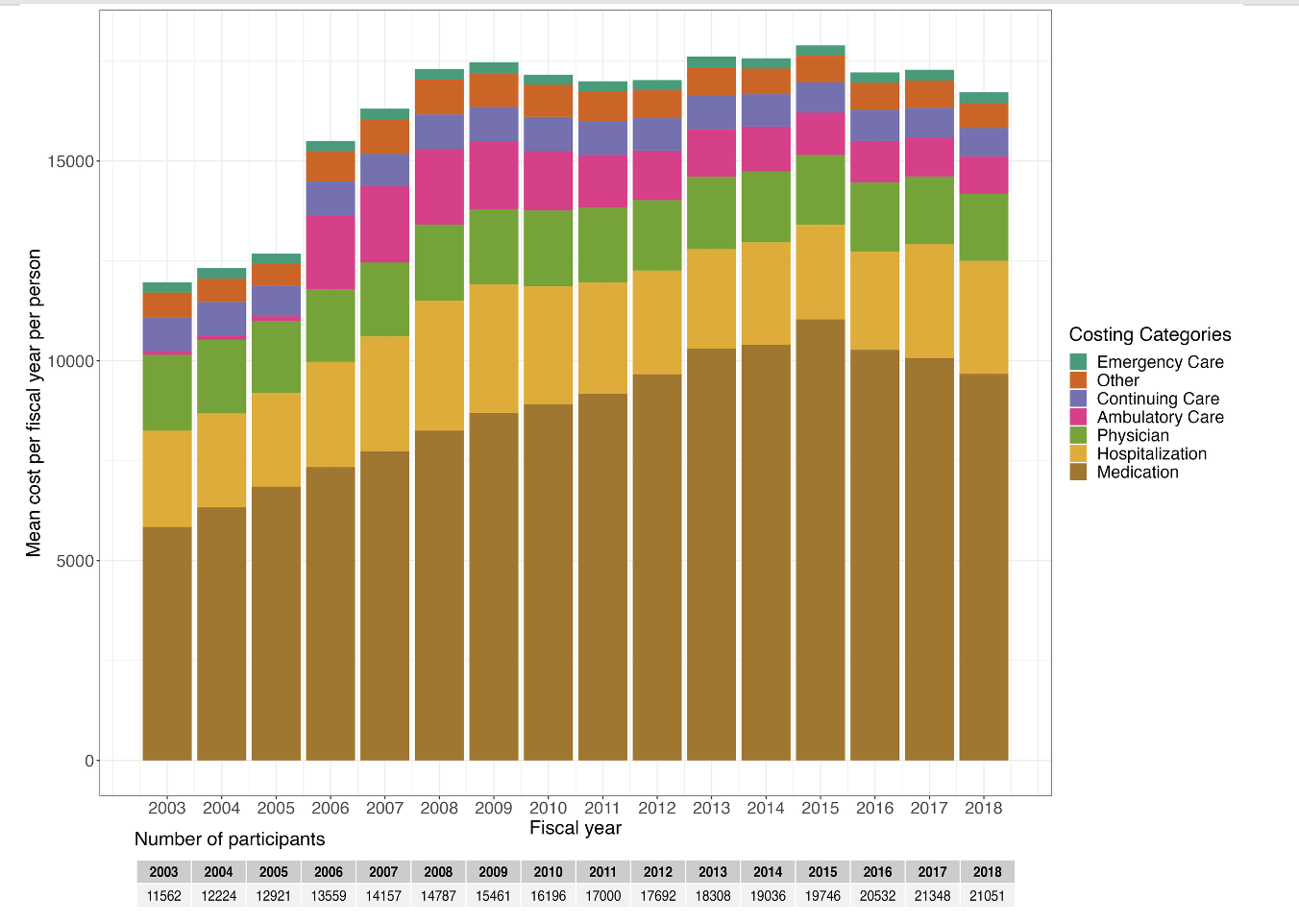
