## Supplementary figures and images for "Trends in Healthcare Costs among People Living with HIV in Ontario, Canada, 2003-2018: Results from a Population-Based Retrospective Cohort Study"

### Appendix C

**Appendix C:** Mean annual direct healthcare costs per person by age category (2025 Canadian Dollars).

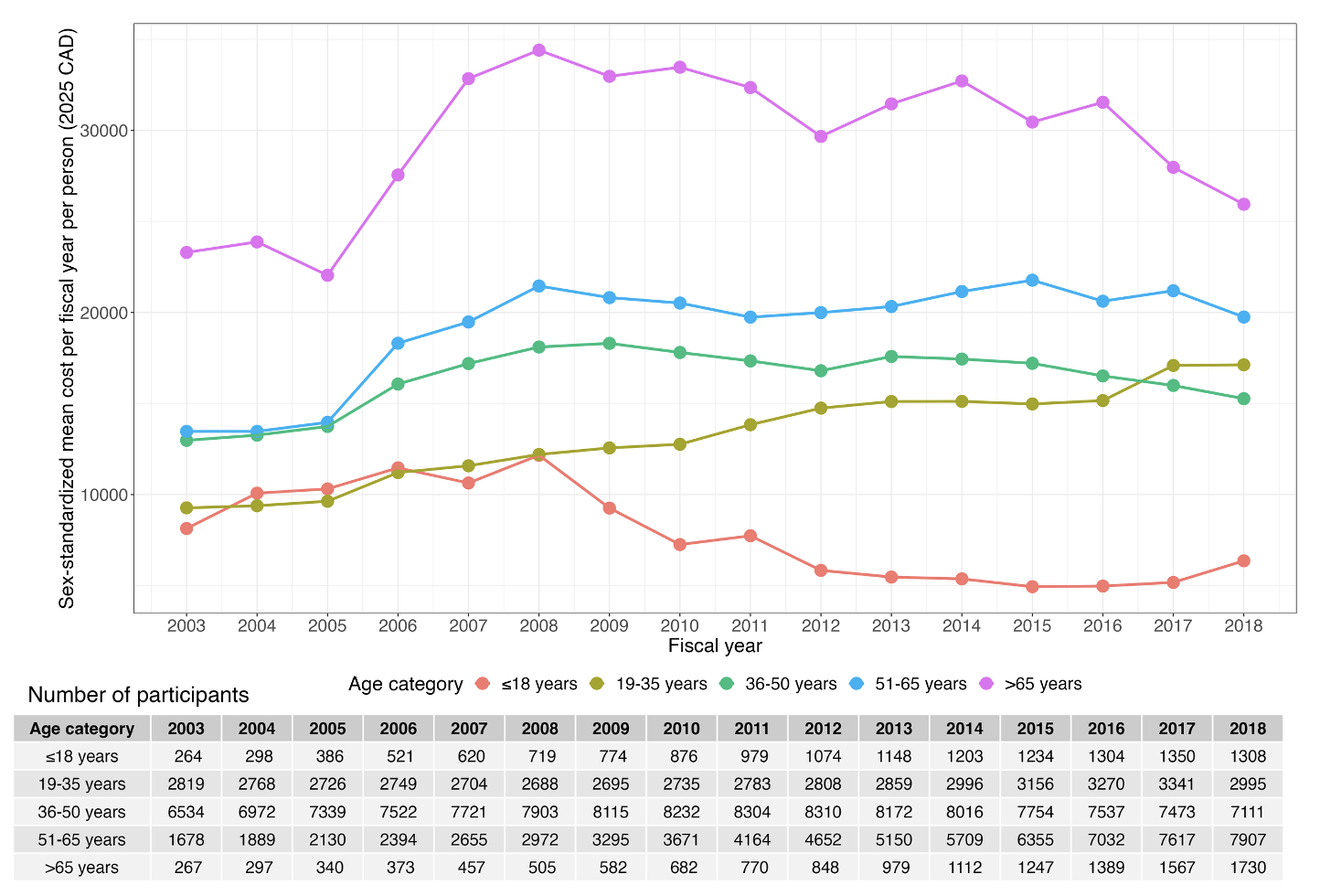

### Appendix D

**Appendix D:** Mean annual direct healthcare costs per person by rurality (2025 Canadian Dollars).

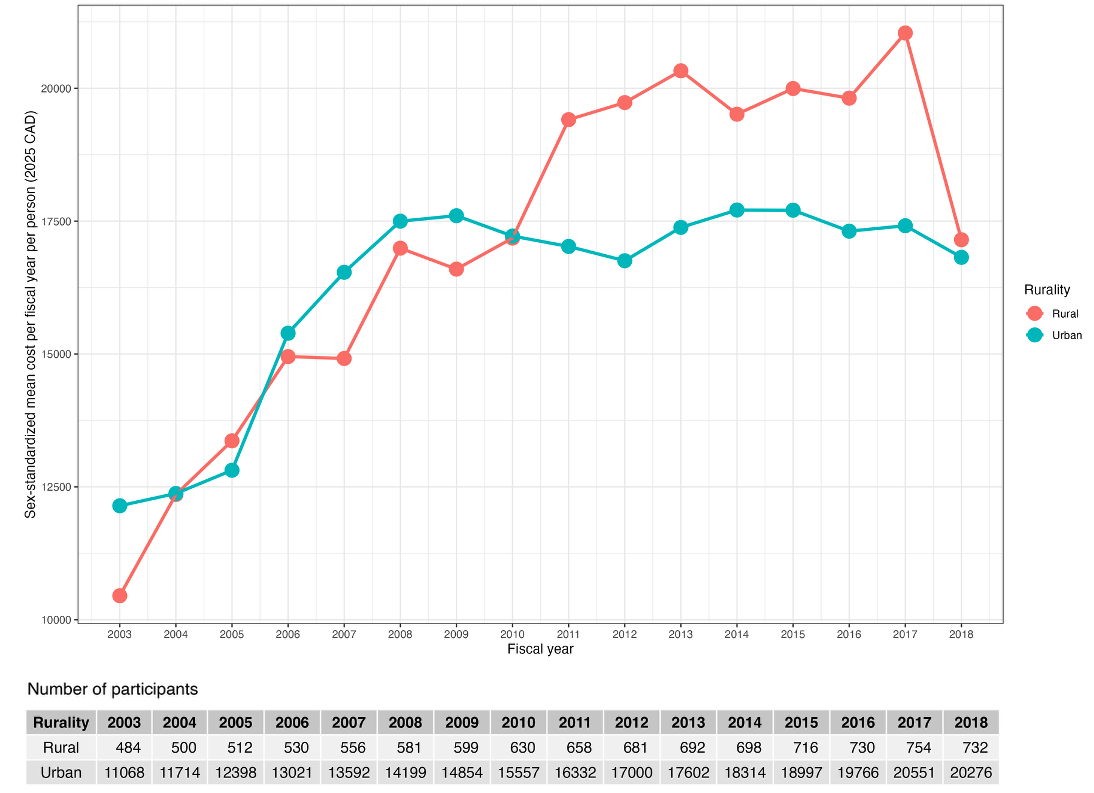
