## Appendix E for "Trends in Healthcare Costs among People Living with HIV in Ontario, Canada, 2003-2018: Results from a Population-Based Retrospective Cohort Study"

**Appendix E:** Mean annual direct healthcare costs per person by immigration status (2025 Canadian Dollars).

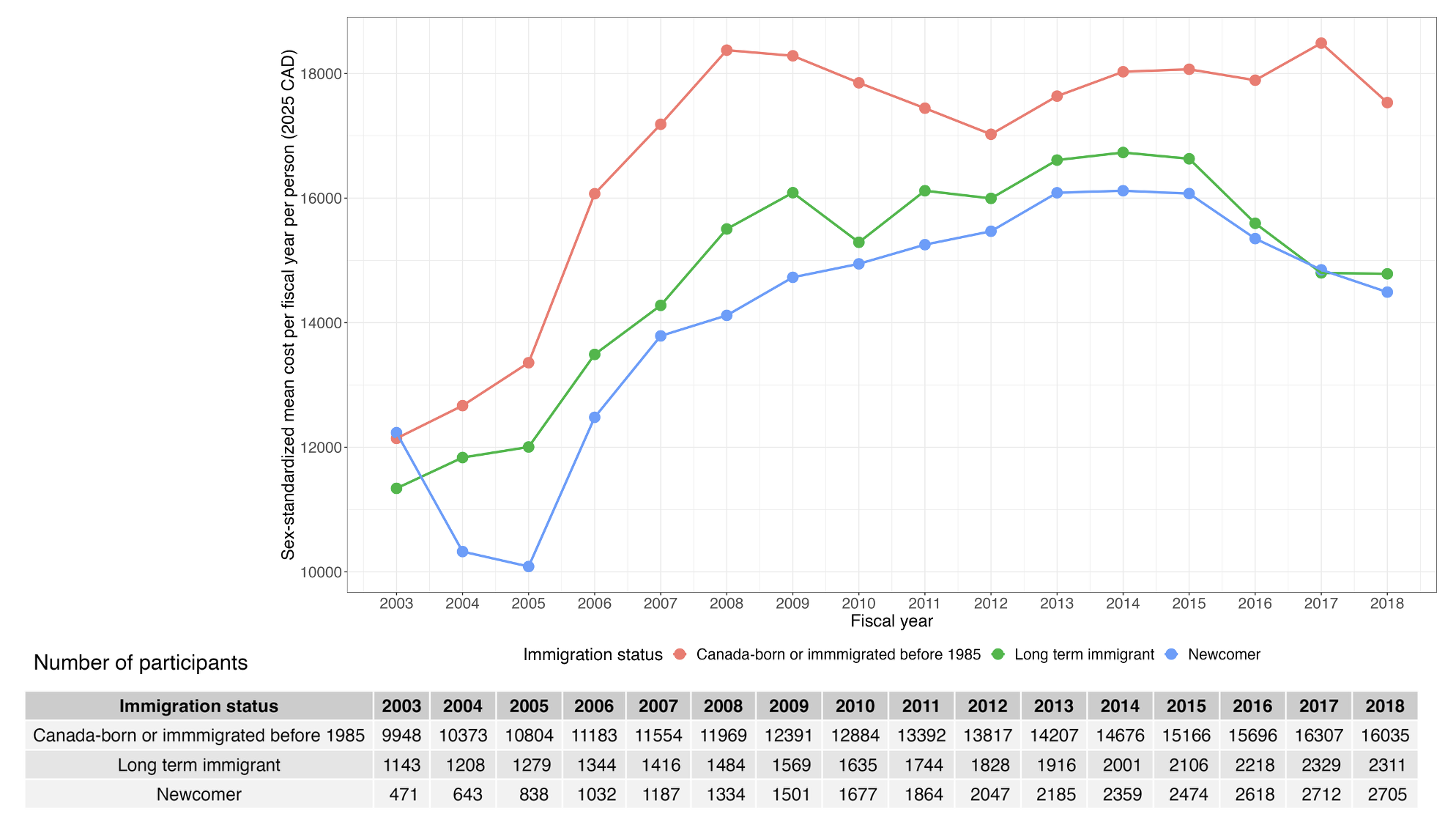

Note: Newcomer refers to people living with HIV who immigrated to Ontario within five years prior to cohort entry.
