## Appendix F for "Trends in Healthcare Costs among People Living with HIV in Ontario, Canada, 2003-2018: Results from a Population-Based Retrospective Cohort Study"

**Appendix F:** Mean annual direct healthcare costs per person by neighbourhood-level income quintile (2025 Canadian Dollars).

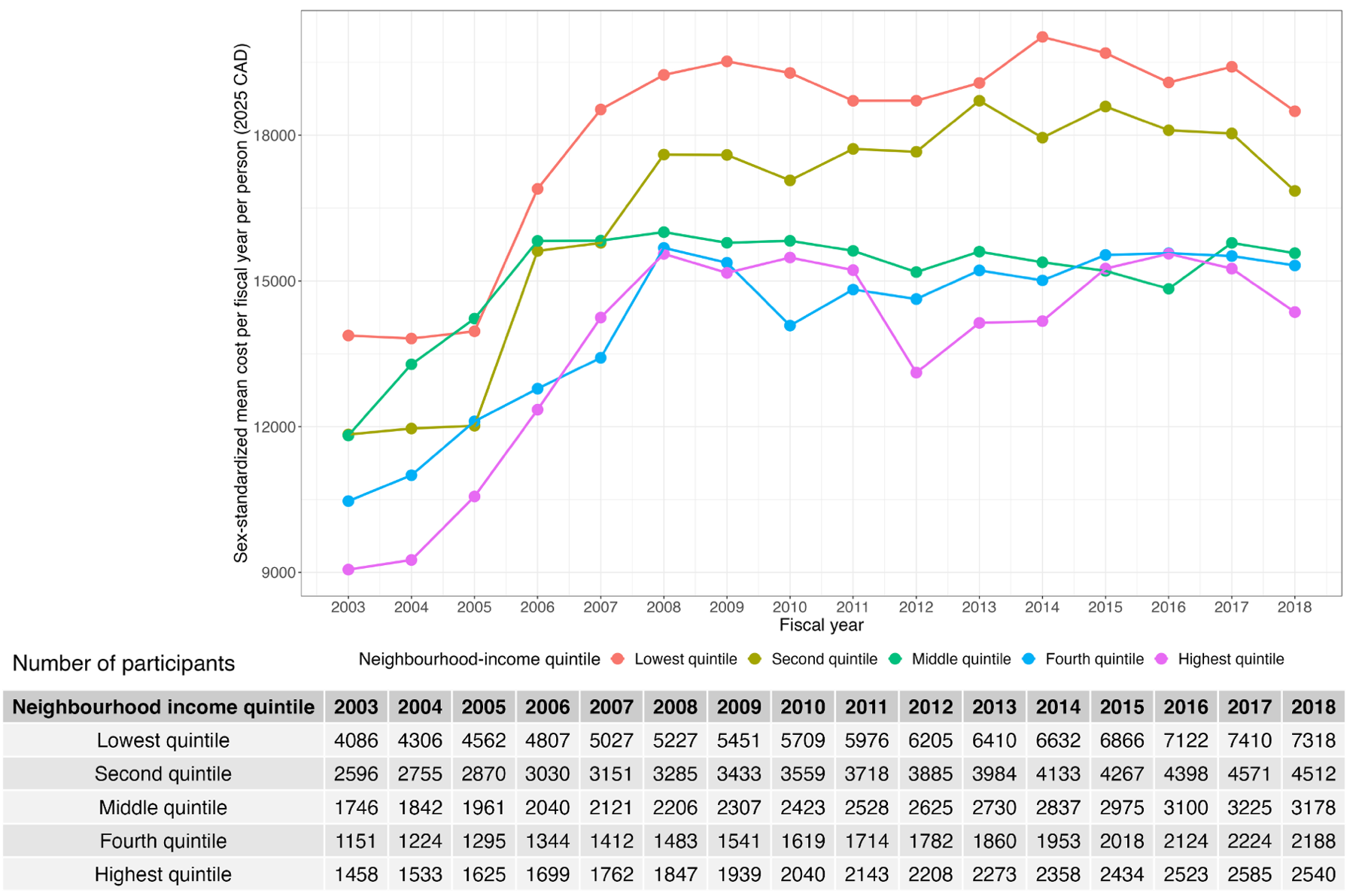

Note: Income quintile has variable cutoff values in each dissemination area (i.e., neighbourhood) in order to take cost of living into account. A census dissemination area being in the highest quintile means it is among the highest 20% of dissemination areas in its city by median household income.
