## Appendix G for "Trends in Healthcare Costs among People Living with HIV in Ontario, Canada, 2003-2018: Results from a Population-Based Retrospective Cohort Study"

**Appendix G:** Mean annual direct healthcare costs per person by ART use ever (2025 Canadian Dollars).

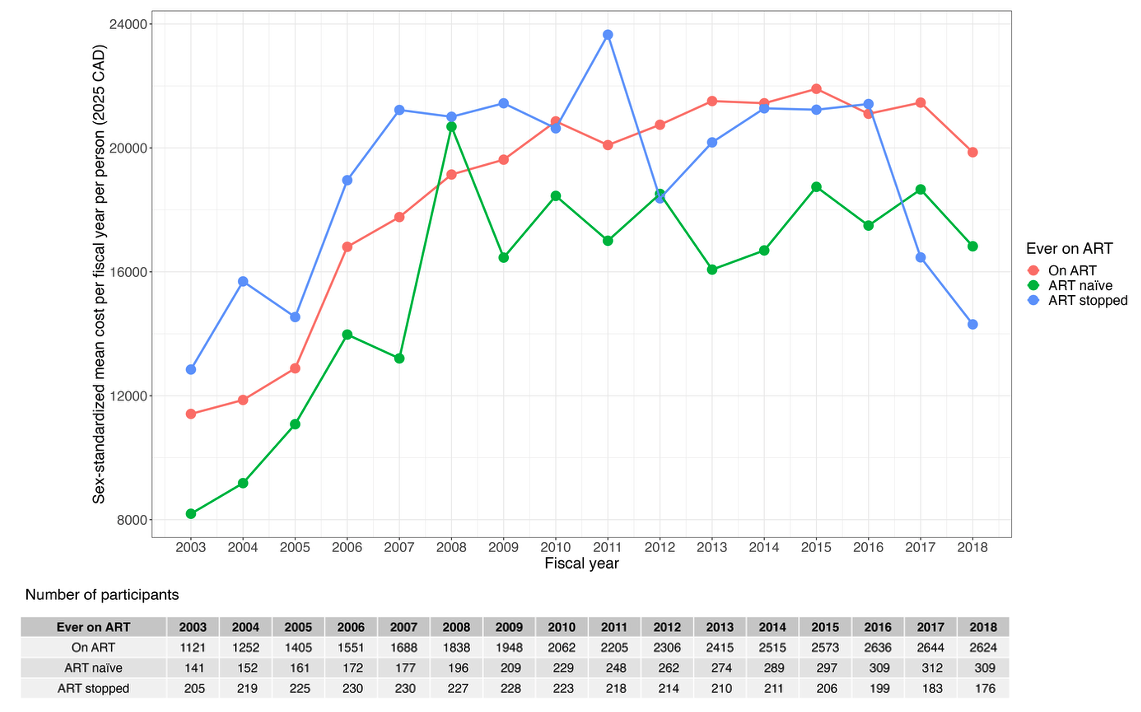

Abbreviations: ART = antiretroviral therapy
