## Appendix H for "Trends in Healthcare Costs among People Living with HIV in Ontario, Canada, 2003-2018: Results from a Population-Based Retrospective Cohort Study"

**Appendix H:** Proportion of mean cost per person per year by costing category and whether individual was ever on ART.

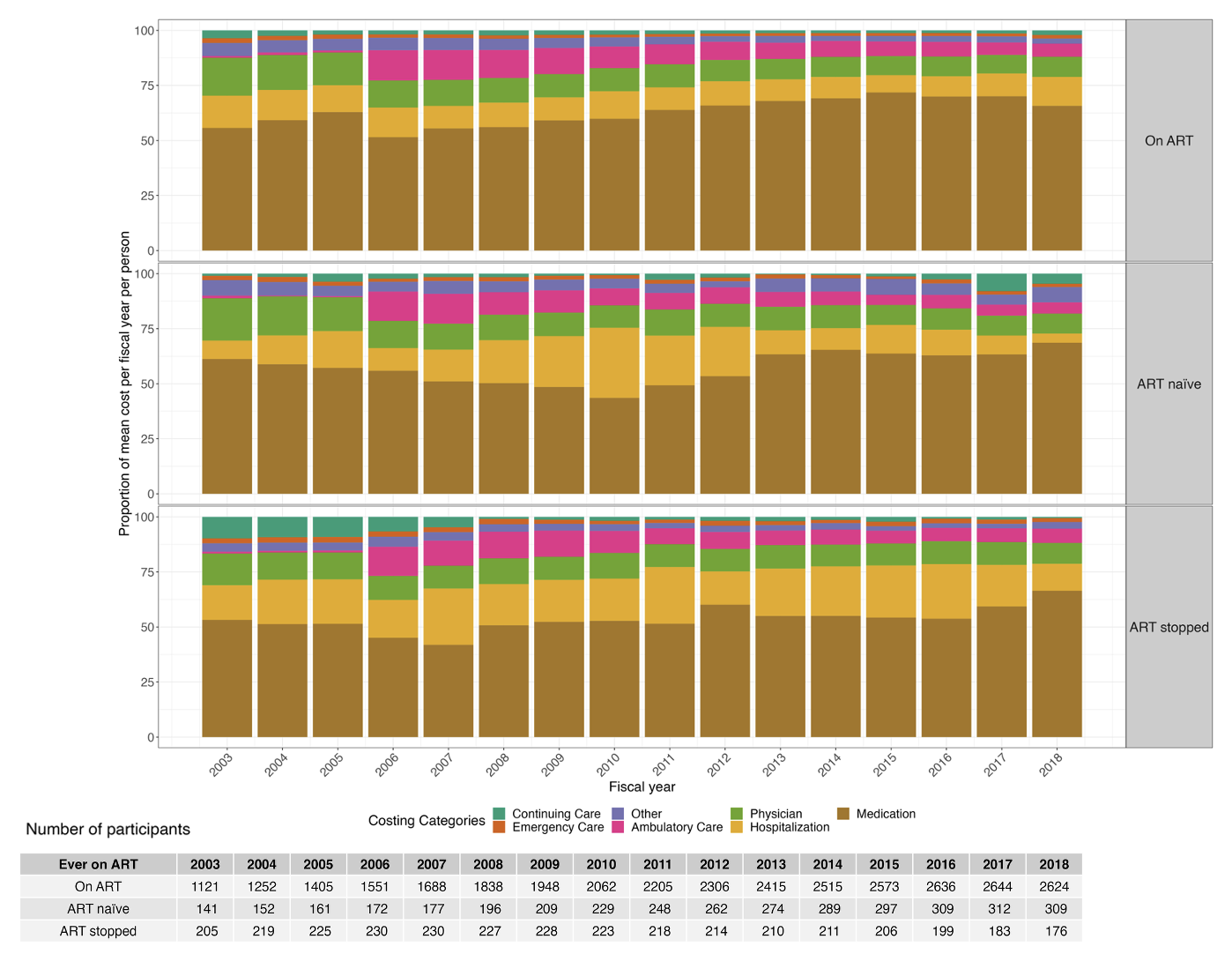

Abbreviations: ART = antiretroviral therapy
